# Diagnostic Utility of Colposcopy and HPV E6/E7 mRNA Testing in Detecting Precancerous and Early Invasive Cervical Lesions among Women Undergoing Cervical Screening in Bangladesh

**DOI:** 10.1101/2025.11.10.25339950

**Authors:** Mohuwa Parvin, Jannatul Ferdous, Farhana Khatoon, Kashfia Binte Quasem, Sumaiya Sadika, Md. Imtiaz Alam

## Abstract

**Background:** Cervical cancer screening aims at detecting precancerous and early invasive lesions to prevent morbidity and mortality. Although colposcopy is a frequently employed visual diagnostic tool with high sensitivity, it has low specificity which often leads to unnecessary biopsies and overtreatment. A possible molecular tool that can enhance the accuracy of the diagnosis is HPV E6/E7 mRNA testing which identifies transcriptionally active viral oncogenes. The study evaluated diagnostic accuracy of colposcopy and HPV E6/E7 mRNA test on women who underwent a cervical screening.

**Methods:** This cross-sectional study included 150 women aged 30-65 years that underwent a cervical screening at a tertiary hospital in Bangladesh. The baseline parameters like parity, menopausal condition and OCP use were noted. The participants underwent colposcopy and HPV E6/E7 mRNA testing, and the histopathology biopsy was the gold standard of lesion grade.

**Findings:** Colposcopy had high sensitivity (94.12%) and very low specificity (8.54) to identify all precancerous and early invasive lesions, which yielded a moderate PPV (46.04%), and accuracy (47.33). Conversely, the E6/E7 mRNA test showed high specificity (93.9%), PPV (78.3%), and low sensitivity (26.5%) and moderate accuracy (63.3%). In the CIN2+ lesions, the sensitivity of colposcopy was very high (90.9 and 98.02), whereas specificity (77.34) and PPV (40.82) scores were low. E6/E7 mRNA test has equal sensitivity (72.7%), good specificity (94.5%), and more accurate (91.3%). The correlation was shown to be significant between E6/E7 mRNA positivity and high-grade lesions that were biopsy-confirmed (p = 0.001), which highlights the clinical relevance of the specified method. It is interesting to note that 15 percent of the women carried high levels of E6/E7 mRNA, which is an indication of transcriptionally active HPV infections.

**Conclusions:** Colposcopy is a very sensitive screening test but it lacks specificity. HPV E6/E7 mRNA has better specificity and accuracy particularly to high grade lesions thus makes it a great complementary or triage test. Combining molecular testing with colposcopy has the potential to enhance screening of cervical cancer by enhancing the accuracy of diagnosis, reducing unnecessary biopsies, and steering to acceptable therapeutic treatment.

## Background

Cervical cancer is one of the most important causes of cancer-related morbidity and mortality amongst women across the globe particularly in the resource-strained regions where screening and early detection initiatives are not easily available [1, 2]. At this point, the natural history of cervical cancer is strongly associated with continued infection with high-risk human papillomavirus (HPV) types, which induce oncogenic alteration in large part via the expression of viral oncoproteins E6 and E7 [3, 4]. These oncoproteins disrupt tumor suppressor pathways leading to cell proliferation and differentiation of low-grade lesions to high grade cervical intraepithelial neoplasia (CIN) and invasive carcinoma [5, 6].

The conventional screening procedures that have been used to detect women at risk with cervical cancer include cytology and colposcopy [7, 8]. The visual method used in screening is called colposcopy that is highly sensitive and not very specific, which leads to unnecessary biopsy and overtreatment [9]. The determination of HPV oncogene expression, especially, E6 and E7 mRNA transcripts, has become a promising approach to enhancing diagnostic accuracy by detecting transcriptionally active infection [10, 11].

Screening programs are done to identify precancerous lesions at early stage so that early management can be done and the development into invasive cancer can be prevented [12]. Popular screening and diagnostic procedures are conventional cytology (Pap smear) and colposcopy. A highly sensitive test, colposcopy which involves the ability to observe the cervix after applying acetic acid, is known to be very sensitive in identifying aberrant epithelial changes [13]. Its sensitivity is, however, often low-quality giving rise to a large number of false positives and unnecessary biopsies [14]. This limitation may cause over-treatment, increased spending on health care and mental suffering on patients.

Recent advances of molecular diagnostics have introduced HPV DNA and mRNA testing as a complement or substitute of the current screening systems [15]. Although HPV DNA tests help to identify that there is a viral genetic material, they do not distinguish between acute infections and those that are transient and do not have carcinogenic ability [16]. Conversely, E6/E7 mRNA transcripts discovery is evidence of transcriptionally active infections, which are more meaningfully associated with high-grade lesion presence and the risk of progression [11]. Consequently, the use of E6/E7 mRNA assays can make cervical cancer screening more specific and able to identify women with clinically severe infections who need further follow-ups or treatment [17].

The sample population in the present investigation covered 150 women aged between 30 and 65 years and undergoing cervical screening and consisted of a group of heterogeneous women with reproductive histories and risk factors. Most of them were multiparous and one out of every four was postmenopausal which would impact cervical epithelial susceptibility and HPV persistence. Moreover, a significant percentage of respondents took the oral contraceptive pill (OCP), which was found to regulate changes in the cervical epithelium and could have been the factor contributing to variations in the dynamics of HPV infections.

The urgent necessity of more reliable and precise screening strategies is the discussed issue that is covered by evaluating the diagnostic performance of colposcopy with HPV E6/E7 mRNA tests. The results illuminated on the complementary roles of eye inspection and molecular tests in identifying precancerous and early invasive lesions of the cervix. The knowledge of advantages and disadvantages of both methods is the key to improving the regimen of cervical cancer screening to eliminate wasteful procedures and, finally, enhance the outcomes of the patient. The fact that the study employed a well-characterized population who came to routine screening can also be seen as an indication that the findings of the study were relevant and applicable to the real-world clinical practice.

## Methods

### Study design and setting

This cross-sectional study was conducted over 1 year (October 2023-September 2024) at Bangladesh Medical University (BMU), Dhaka, Bangladesh, in the Colposcopy Clinic of the National Centre of Cervical and Breast Cancer Screening and Training.

### Study population

One hundred and fifty women aged between 30 and 65 years that were VIA-positive, had abnormal cytology or positive for HPV DNA on routine screening were included. Pregnant women, those who had undergone a hysterectomy in the past, or whose cervix had received LEEP or conization, known cases of aggressive cancer, and those who did not give their consent were excluded

### Collection and Processing of samples

After the confirmation of the eligibility, each subject received an extensive gynecological examination and colposcopy with the help of the traditional visualizing tools. A sterile cytobrush was used to collect cervical exfoliated cells and without delay transfer them to Microtubes, RNase-free and Buffer RL of the Macro & Micro-Test Viral DNA/RNA Kit (Catalog No: HWTS-3020-50bd). The samples obtained were put in ice and forwarded to the Department of Virology to extract RNA and conduct molecular tests.

### RNA Extraction Procedure

The Macro and Micro-Test Viral DNA/RNA Kit was used to extract total RNA, using the standard protocol mentioned on the kit, out of cervical cells. In short, cervical samples (up to 20 mg tissue equivalent) were pre-homogenized in 300 -mercaptoethanol in 300 ûL Buffer RL to inactivate RNases. Then, 10 uL of Proteinase K solution was placed and incubated at 56 o C throughout 10-20 minutes to digest protein. Subsequently, an absolute volume of ethanol (0.5 vol) was put in the lysate and swirled. The lysate was added to an RNase-free spin column (CR3) and centrifuged at 12000 rpm within a period of 30-60 seconds so that the RNA attaches itself to the silica membrane. Buffers RW I and RW II were used twice to wash the column in order to eliminate impurities. As a control to remove the DNA contamination, 10 μL of DNase I stock solution combined with 70 μL of Buffer RDD was introduced directly into the column and incubated with it over 15 minutes at room temperature. Following the washing, the total RNA was washed with 30 μL of the RNase-free water and kept at the temperature of -70 C prior to PCR analysis.

### cDNA Synthesis and E6/E7 mRNA Detection

HPV E6/E7 mRNA with a one-step RT-PCR method was detected in the QuantStudio 5 Real-Time PCR System (Applied Biosystems Thermo Fisher Scientific, USA) with the transcript of high-risk HPV type 16, 18, 31, 33, and 45. Every reaction mixture was composed of 10 μL extracted RNA template, 1x One-Step RT-PCR Master Mix, gene-specific primers and fluorescent probes, and RNase-free water (25 μL). The thermal cycling parameters involved reverse transcription at 50 o C of 15 minutes, initial denaturation temperature at 95 o C of 2 min and 40 cycles of 95 o C of 15 s and 60 o C of 1 min. QuantStudio Design and Analysis Software v1.5 was used in analyzing the real-time fluorescence data. Samples that had amplification signals that hit the fluorescence threshold at least 40 times were taken as positive in terms of HPV E6/E7 mRNA expression.

### Histopathological Examination

At the same time, the cervical punch samples were obtained with the help of a colposcopic examination of the suspicious transformation-zone sites and examined in the Department of Pathology. The tissue was fixed using 10% neutral buffered formalin, paraffin embedded, sectioned at 3-5 μm and stained with hematoxylin and eosin stain (H&E). A histopathologist assessed each of the slides and determined it to be CIN 1, CIN 2, CIN 3, or invasive cancer according to the WHO classification of 2014. The diagnostic gold standard to assess the accuracy of the tests was the results of histopathology.

### Data Analysis and Statistical Data Management

Every demographic, clinical and laboratory data was entered into a standardized case record form and analyzed in SPSS version 26.0 (IBM Corp., Armonk, NY). Categorical variables were formulated in the form of frequencies and percentages and continuous variables were formulated in terms of mean standard deviation. Using histopathology as a reference standard, the sensitivity, specificity, positive predictive value (PPV) and negative predictive value (NPV) and overall accuracy of colposcopy and HPV E6/E7 mRNA testing were established. Chi-square or Fisher exact test was used to compare categorical variables where p-values of less than 0.05 were considered significant.

### Ethical Considerations

Ethical clearance was approved by Institutional Review Board (IRB) of Bangladesh Medical University (IRB registration number-4628 and date-01.10.2023). Informed permission to participate in the study was given in writing before participation and the objectives, procedures, risks, and benefits of the study were explained to all the participants.

## Results

### Baseline Characteristics of the Study Participants

This was a study that involved 150 women between the ages of 30 and 65 years. The largest participation rate (45.3%) was observed in the age group of 30-39 years, then 40-49 years (30.7%), and 50 and above age (24.0). Most of the women (87.3) were multiparous with over a quarter (27.3) of them being postmenopausal. The duration of marriage among participants was 21.3 with a standard deviation of 5.6 indicating that the participants engaged in a lot of sex life. Irrespective of them, these characteristics make up a representative sample of women who screening of their neck occurs in this population (Table 1).

**Table 1:** Baseline Characteristics of the Study Participants (n = 150)

| Characteristics | Frequency (%) |
| --- | --- |
| **Age (years)** |  |
| 30–39 | 68 (45.3) |
| 40–49 | 46 (30.7) |
| ≥50 | 36 (24.0) |
| **Parity** |  |
| Multiparous | 131 (87.3) |
| **Menopausal status** |  |
| Postmenopausal | 41 (27.3) |
| **Duration of marriage (years)** | Mean ± SD = 21.3 ± 5.6 |

### OCP Use among Study Participants

Out of the 150 women involved in the study, 62.0% (n = 93) were the women who were using the oral contraceptive pills (OCPs), and 38.0% (n = 57) were not on the contraceptives. The population of OCP users is dominant in the study population represented by the distribution of OCP use in a donut chart provided in figure 1. This observation indicates a quite a large percentage of female contraception use in the study setting of cervical screening of women (Figure 1).

**Figure 1:**
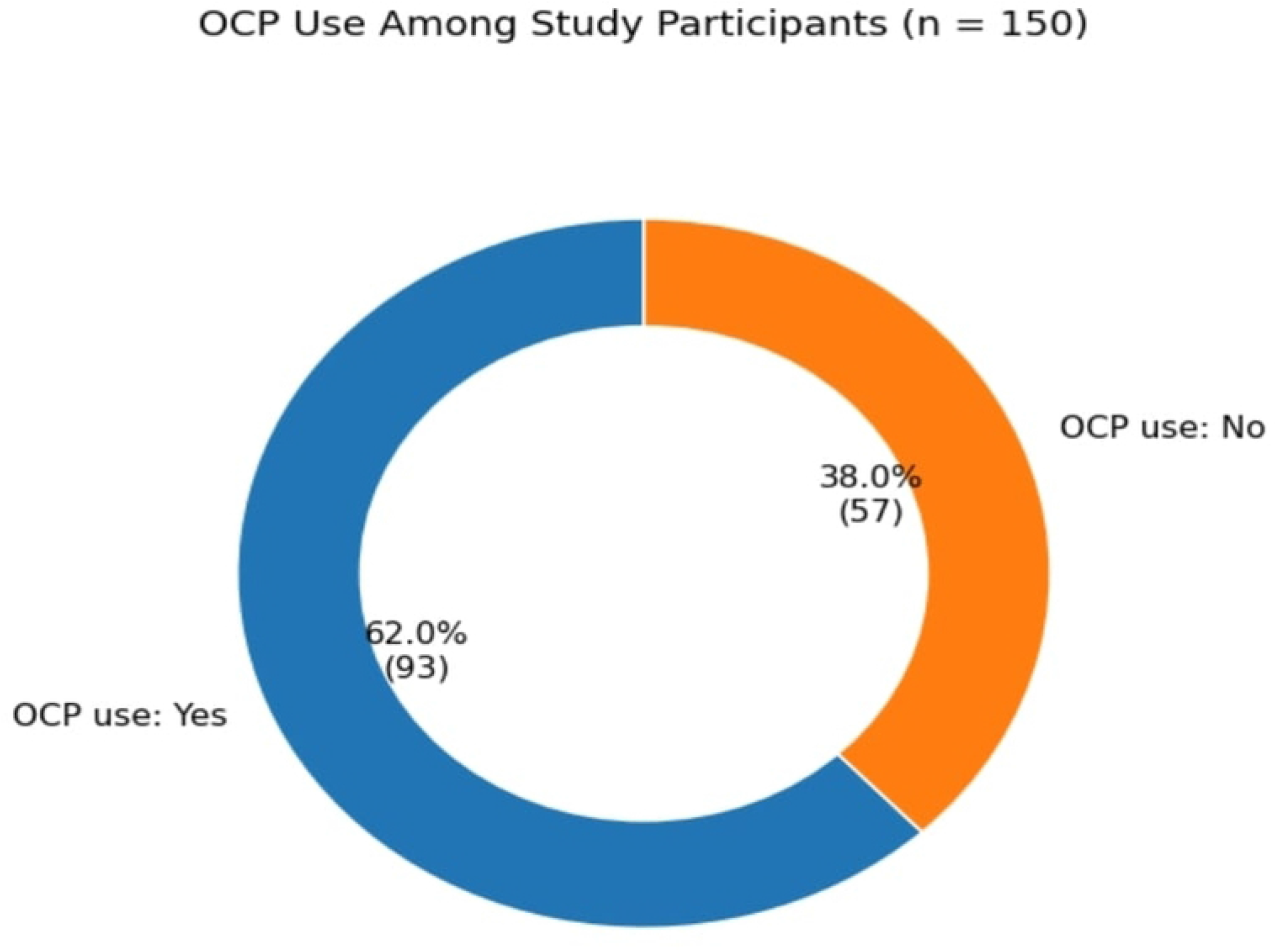
Donut chart showing the distribution of oral contraceptive pill (OCP) use among study participants (n = 150)

### Diagnostic Performance of Colposcopy and E6/E7 mRNA Testing for Precancerous and Early Invasive Lesions

Diagnostic Performance of Colposcopy and E6/E7 mRNA Testing to Precancerous and Early Invasive Lesions. Colposcopy was very sensitive (94.12% - when all grades of precancerous and early invasive lesions of the cervix were evaluated) but very low in specificity (8.54%). By comparison, the E6/E7 mRNA test was significantly more specific (93.9%), positive predictive (78.3%), and intermediate (63.3). Although the E6/E7 mRNA test had the lowest sensitivity (26.5%), it was much more effective in identifying true negative cases, and it had a reduced number of false positives compared to colposcopy. This paper highlights that genetic testing has a possibility to complement visual tests in the screening of cervical cancer (Table 2).

**Table 2:**
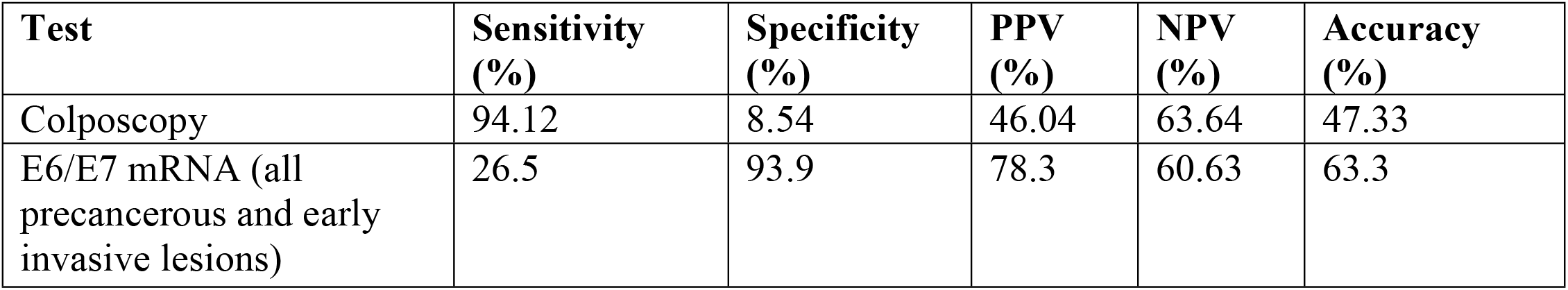
Diagnostic Performance of Colposcopy and E6/E7 mRNA Testing for Precancerous and Early Invasive Cervical Lesions.

| **Test** | **Sensitivity (%)** | **Specificity (%)** | **PPV (%)** | **NPV (%)** | **Accuracy (%)** |
| --- | --- | --- | --- | --- | --- |
| Colposcopy | 94.12 | 8.54 | 46.04 | 63.64 | 47.33 |
| E6/E7 mRNA (all precancerous and early invasive lesions) | 26.5 | 93.9 | 78.3 | 60.63 | 63.3 |

### Association between Colposcopy, E6/E7 mRNA Expression, and Histopathological Findings for CIN2+ Lesions

In high grade lesions (CIN2+), there were marked correlations among diagnostic modalities. Though precancerous changes were apparent on colposcopy in 20 of 139 cases (14.4) with coexisting E6/E7 mRNA positivity, three of eleven women (27.3) who appeared normal on colposcopy were E6/E7 mRNA positive and this proved that the disease at a molecular level was not visible by visualization (p = 0.376). Colposcopy and biopsy results were found to have strong correlation (p = 0.001), which validates the argument that histopathology remains the gold standard with regard to lesion grading. Notably, there was a considerably high degree of significance (p = 0.001) between the presence of E6/E7 mRNA and biopsy-validated CIN2+ lesions with 69.6 percent of mRNA positive sample results indicating high grade disease. This supports the diagnostic power of mRNA assay to identify clinically significant abnormalities (Table 3).

**Table 3:** Association between Colposcopy, E6/E7 mRNA Expression, and Histopathology Findings for CIN2+ Lesions.

| Comparison | CIN2 and Higher n (%) | < CIN2 n (%) | p-value |
| --- | --- | --- | --- |
| **Colposcopy vs. E6/E7 mRNA** | Pre-cancerous (20/139; 14.4%) vs. Normal (3/11; 27.3%) | — | 0.376 |
| **Colposcopy vs. Biopsy** | 20 (40.8%) vs. 2 (1.98%) | 29 (59.2%) vs. 99 (98.02%) | 0.001 |
| **E6/E7 mRNA vs. Biopsy** | 16 (69.6%) vs. 6 (4.7%) | 7 (30.4%) vs. 121 (95.3%) | 0.001 |

### Comparative Diagnostic Performance for CIN2 and Higher Lesions

Colposcopy was sensitive (90.9%), had a negative projected value (NPV) of 98.02 percent of high grade cervical intraepithelial lesions (CIN2+), but low specificity (77.34%) with low PPV (40.82%). E6/E7 mRNA test on the other hand was highly specific (94.5) and accurate (91.3) with a balanced sensitivity (72.7) and NPV (95.3). The results indicate that, although colposcopy is a very sensitive test in the detection of lesions, E6/E7 mRNA assay is more precise and diagnostic, and should be used as a triage method in the identification of high-grade cervical disease and the prevention of unwarranted interventions (Table 4).

**Table 4:** Comparative Diagnostic Performance of Colposcopy and E6/E7 mRNA Testing for CIN2+ Lesions.

| Test | Sensitivity (%) | Specificity (%) | PPV (%) | NPV (%) | Accuracy (%) |
| --- | --- | --- | --- | --- | --- |
| Colposcopy | 90.9 | 77.34 | 40.82 | 98.02 | 79.3 |
| E6/E7 mRNA (CIN2 and higher) | 72.7 | 94.5 | 69.56 | 95.3 | 91.3 |

### Expression Pattern of HPV E6/E7 mRNA

The overexpression of HPV E6/E7 mRNA was detected in 23 (15%) of 150 women who were tested and 127 (85) women did not have any detectable E6/E7 transcripts. The high mRNA-negative findings reveal that most of the screen-positive women had no transcriptionally active high-risk HPV infections when they had the test. Such a molecular distribution demonstrates that a very small fraction of the women are found to be transcriptionally active with viral oncogenes, which is associated with a greater predisposition to form high-grade cervical lesions (CIN2+). Consequently, E6/E7 mRNA testing may become a more precise triaging intervention to designate high-risk women among women with abnormal cytology or women with positive screening outcomes (Figure 2).

**Figure 2:**
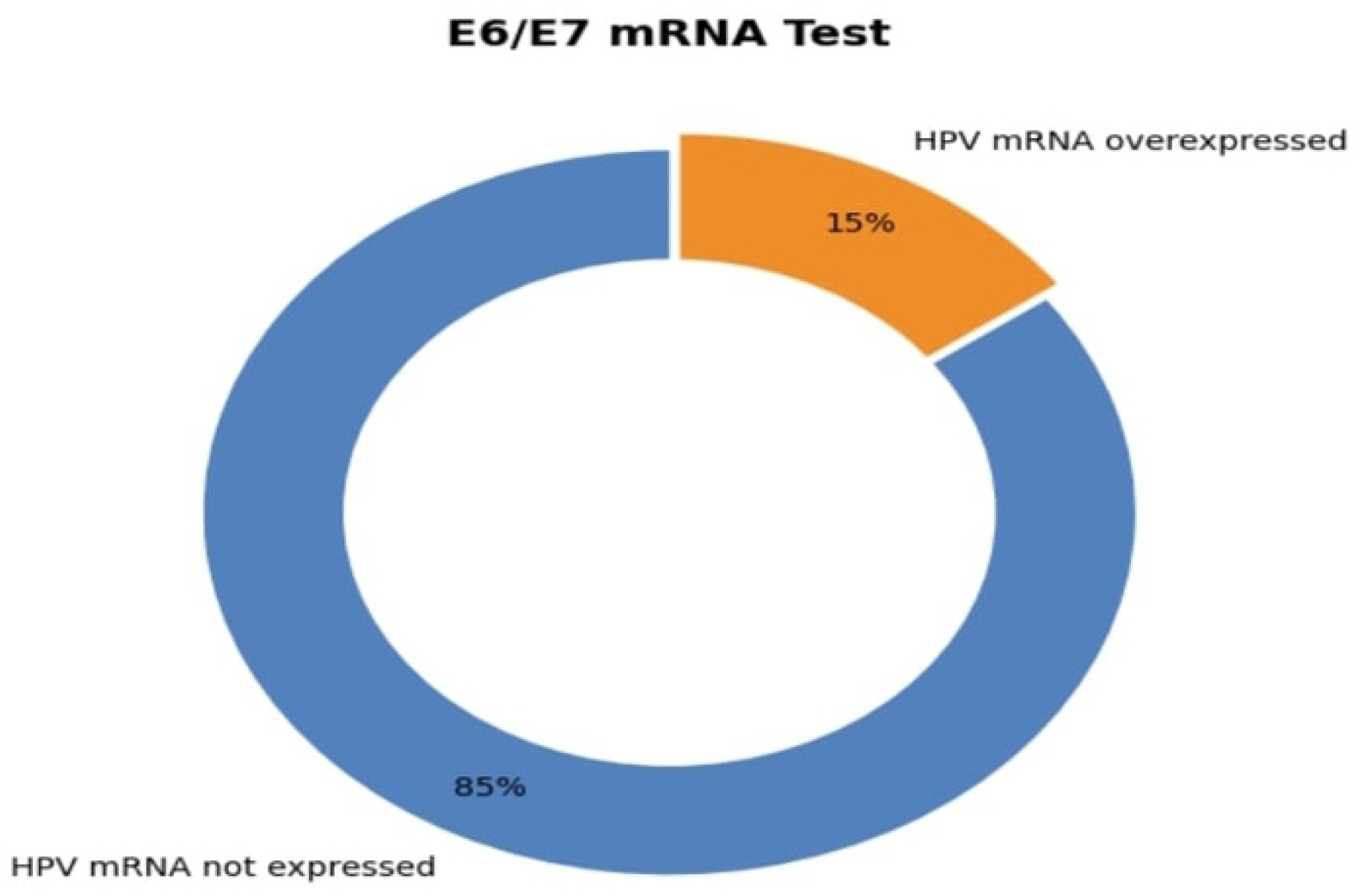
Distribution of Participants According to Overexpression of E6/E7 mRNA.

## Discussion

The results of the study are very important to the diagnosis of precancerous and early invasive cervical lesions in colposcopy and HPV E6/E7 mRNA tests. Colposcopy demonstrated a very high sensitivity (94.12) in all grades of lesions, and has been found useful as the initial visual screening tool. But, its extremely low specificity (8.54%), means that there is a high likelihood that it will have false positives, and thus lead to falsely positive biopsies, and patient suffering [18]. This fact shows the necessity of additional testing with the focus on the specificity. When the lesion grades were considered overall, the E6/E7 mRNA assay was significantly more specific (93.9%), and positive predictive value (78.3%), but with lower sensitivity (26.5%). This demonstrates that even though the mRNA test might be incapable of detecting all lesions, it is more precise in confirming the presence of the disease and reducing false positives. The oncogene expression in E6/E7 transcripts was detected on the molecular level and is more likely to be associated with the clinical severe lesions [19,20].

Colposcopy was found to be useful in the detection of the disease when it targeted high-grade lesions (CIN2+) because it was characterized by a high sensitivity (90.9%), as well as a high negative predictive value (98.02%). Nonetheless, its specificity (77.34) and positive predictive value (40.82) were moderate indicating some over diagnosis. The sensitivity (72.7) of the E6/E7 mRNA test was balanced, specificity (94.5) was higher, and accuracy (91.3) of CIN2+ lesions was high. This balance serves to justify its use as a useful triage instrument of identifying women at a high-risk of high-grade illness, who may inform more specific biopsy and treatment choices [21].

Clinical application of the assay is supported by the high correlation levels between E6/E7 mRNA positivity and biopsy-detected CIN2+ lesions (p = 0.001). It is important to note that, out of the normal colposcopic outcome, there are certain women who were positive in F6/E7 mRNA as a sign that the molly-code can detect an early or occult disease that cannot be seen on colposcopy [22]. The free association between ocular and molecular diagnosis can enhance the effectiveness of screening in general [23].

HPV E6/E7 mRNA pattern of expression, which is only overexpressed in 15 percent of women, indicates that active transcription of high-risk HPV is rare in screen-positive women. This molecular stratification can guide clinicians to concentrate their efforts on individuals that face the highest likelihood of benefiting through intervention reducing overtreatment of women who have temporary or inactive illnesses [24].

It was found that distinctly there is a significant disparity in specificity between the E6/E7 mRNA test and colposcopy, which presents a very important challenge of cervical cancer screening; between sensitivity to ensure that no potential lesions are missed and specificity to reduce over diagnosis. Colposcopy is very sensitive to find most lesions, but it has low specificity that can result in unnecessary biopsies and increased medical expenses [25, 26]. The fact that the E6/E7 mRNA assay can differentiate between the true positive and the false positive is a strength of the test in treating a patient since it is able to identify individuals who have transcriptionally active HPV infections that are more likely to cause the progression.

Overall, even though colposcopy is a sensitive and readily available test in identifying lesions of the cervix, its low specificity limits its application in creating a one-test system. Specificity and diagnostic accuracy can be enhanced by incorporating HPV E6/E7 mRNA testing in high-grade lesions, meaning that it can be incorporated into cervical cancer screening algorithms as a screening (triage) test. This is a combination strategy that can optimize the use of resources, limit unwarranted treatment, and eventually improve patient outcomes during cervical cancer preventive programs.

## Strengths and Limitations

The study has a clear and representative sample of women aged between 30 and 65 years and had gone through cervical screening, which enhances the applicability of the findings to the actual clinical practice. When there is a complete comparison of colposcopy and HPV E6/E7 mRNA testing to the gold standard of histopathology, some critical data is obtained on the diagnostic strengths and limitations of the visual and molecular studies. Moreover, the provision of the specific diagnostic data of all lesion grades and high-grade CIN2+ lesions facilitates a finer perspective on the results of tests. Nonetheless, the small sample size of the study can reduce the statistical capability and accuracy of the results, particularly to the less common high-grade lesions.

Its cross-sectional nature prevents the examination of long-term consequences like lesions progression and there is the risk of selection bias, since the study involved women who already underwent screening programs, which are not representative of all at-risk groups. Moreover, the fact that the sensitivity of the E6/E7 mRNA test is low in all the grades of lesions demonstrates that this screening method might not be suitable as a single test. Lastly, the paper fails to respond to the issue of cost-effectiveness, which is essential in the real implementation of molecular testing in various healthcare institutions

## Conclusion

In this study, it was established that colposcopy is a very sensitive method of detection of precancerous lesions and early invasive lesions of the cervix, although it has low specificity; hence, it has a high number of false positives. The testing of HPV E6/E7 mRNA is more specific and accurate particularly in detection of high-grade lesions of CIN2+ as this test detects transcriptionally active viral oncogenes that is linked with clinically severe illness. Molecular tests along with colposcopy would be useful in enhancing diagnosis, avoiding unnecessary biopsy, and better management of patients in cervical cancer screening initiatives. More and larger cohort and longitudinal follow-up studies are required in the future to confirm these results and examine the cost-effectiveness and feasibility of integrating E6/E7 mRNA test in routine clinical practice.

## Data Availability

10.6084/m9.figshare.30572345

https://10.6084/m9.figshare.30572345

## Declarations

### Data sharing statement

All data relevant to the study are accessible in <u>10.6084/m9.figshare.30572345</u>

